# Phase Ib Study of Enzalutamide with Venetoclax in Patients with Metastatic Castration-Resistant Prostate Cancer

**DOI:** 10.1101/2025.04.22.25326208

**Authors:** Stuthi Perimbeti, Anmbreen Jamroze, Dharmesh Gopalakrishnan, Rohit Jain, Changchuan Jiang, Julianne L Holleran, Robert A. Parise, Robert Bies, David Quinn, Kristopher Attwood, Xiaozhuo Liu, Jason S. Kirk, Jan H. Beumer, Dean G. Tang, Gurkamal Chatta

## Abstract

**Purpose:** Castration and enzalutamide induce BCL-2 to drive therapy resistance in prostate cancer (PCa). We conducted a phase Ib trial to test that metastatic castration-resistant PCa (mCRPC) can be effectively targeted by combining enzalutamide with the BCL-2 inhibitor venetoclax.

**Experimental Design:** This phase Ib single-arm trial of enzalutamide (160 mg/d) with venetoclax in patients with progressive mCRPC assessed dose-limiting toxicity (DLT), maximum tolerated dose (MTD) and recommended phase 2 dose (RP2D). Three dose levels (DL) of venetoclax (DL1 400 mg/d; DL2 600 mg/d; and DL3 800 mg) were evaluated using a 3+3 design. We also analyzed enzalutamide and venetoclax pharmacokinetics and conducted pharmacodynamic studies in peripheral blood mononuclear cells (PBMCs) to determine the impact of venetoclax on BCL-2 expression.

**Results:** A total of 10 patients were enrolled across 3 DL and no DLT was observed. Mean duration on treatment was 29 weeks (range: 8-140 weeks). Treatment-related adverse events (TRAEs) were mostly grade 1-2, and Grade 3 TRAEs included hypertension (20%), fatigue (10%), and thrombocytopenia (10%). 1/10 (10%) attained PSA50 response and 4/10 (40%) had stable disease. Estimated median overall survival (OS) was 19 months (95% CI 5-28 months) and median time to next systemic therapy (TNST) was 5 months (95% CI 1-35 months). Pharmacokinetic results revealed sub-therapeutic plasma levels of venetoclax. Pharmacodynamic studies demonstrated that venetoclax enhanced BCL-2β generation and promoted BCL-2 degradation.

**Conclusions:** Enzalutamide with venetoclax has an acceptable toxicity profile in patients with mCRPC. Despite sub-therapeutic venetoclax levels, the treatment elicited pharmacodynamic and clinical response in a subset of patients.

Clinical trial ID: NCT03751436

**Translational Relevance:** There are limited treatment options for mCRPC patients. Castration and androgen receptor (AR) targeted therapies cause significant cellular plasticity and exacerbate AR heterogeneity in mCRPC resulting in emergence of AR-positive (AR^+^) and AR-negative (AR^−^) PCa progenitor cells and metastatic foci. While AR^+^ mCRPC can be targeted by AR pathway inhibitors (ARPIs) such as enzalutamide, AR^−^ mCRPC cannot. Based on our recent findings that castration and ARPIs upregulate BCL-2 in both AR^+^ and AR^−^ CRPC models to drive castration resistance, here we conduct a prospective phase Ib clinical study to test that the heterogeneous AR^+^ and AR^−^ mCRPC can be holistically targeted by combining enzalutamide with the BCL-2 inhibitor venetoclax.

## Introduction

More than 35,000 American men are estimated to die from metastatic castration-resistant prostate cancer (mCRPC) in 2025 (1). Most primary prostate cancer (PCa) patients are treated and cured with surgery and/or radiation therapy. Patients with locally advanced or recurrent disease are treated with androgen deprivation therapy (ADT), which blocks testicular androgen production, often in combination with next generation of androgen receptor (AR) pathway inhibitors (ARPIs) such as enzalutamide and abiraterone acetate or with docetaxel-based chemotherapy (2–4). Unfortunately, despite initial response, most patients progress after ∼18-24 months and develop mCRPC. Treatment options for patients with refractory mCRPC are limited. Recent advances have introduced new agents for mCRPC, but questions about treatment sequencing and individualized approaches persist, and with a median survival of about 3 years, mCRPC is invariably fatal (3,4).

Numerous mechanisms have been proposed to explain PCa resistance to ADT and ARPIs, most of which are centered on alterations of AR itself including *AR* genomic amplifications and expression of constitutively active AR splice variants that lack the C-terminal ligand-binding domain (5). One under-appreciated and under-studied mechanism underlying ARPI resistance and CRPC development is mediated by heterogeneity in AR expression and plasticity in PCa cells (6–14). In fact, AR protein expression is heterogeneous in untreated primary tumors, with both AR^+^ and AR^−/lo^ PCa cells co-existing (6–8,14). AR heterogeneity becomes more accentuated in metastases and mCRPC (8–11,13). Recent studies have put this AR heterogeneity in the context of tumor response to ADT/ARPIs and other therapies - although the AR^+^ PCa responds to enzalutamide, AR^−/lo^ PCa and CRPC are inherently resistant to ARPIs (7,8,10,11,13). Because patient primary tumors and CRPC contain both AR^+^ and AR^−/lo^ PCa cells/clones and because ARPIs induce plasticity in PCa cells (by reprogramming AR^+^ PCa cells into AR^−/lo^ cells), it has become clear that both populations of PCa cells need to be therapeutically targeted (12–14).

We have reported preferential expression of the prosurvival and stemness factor BCL-2 in therapy-resistant AR^−/lo^ PCa stem cells (7,8). Our recent xenograft modeling has demonstrated that castration and enzalutamide upregulate BCL-2 to drive ADT/ARPI resistance (11), nominating BCL-2 as a therapeutic target of CRPC. In support, combination of enzalutamide with the BCL-2 inhibitor ABT-199 inhibited the emergence of enzalutamide-resistant AR^+/hi^ LNCaP-CRPC by 75%, and ABT-199 alone (but not enzalutamide) greatly inhibited the growth of AR^−/lo^ LAPC9-CRPC (11). Importantly, multiple studies (15–21) from others also support BCL-2 as a CRPC target. Unfortunately, early clinical trials targeting BCL-2 have failed, mainly due to the use of non-specific and ineffective BCL-2 inhibitors (22–25). Although initially promising, early BCL-2 inhibitors failed to show sufficient clinical activity for leukemia and were never approved by the FDA for use in patients (26–28). Second and third generation BCL-2 inhibitors have subsequently been developed, with significantly increased specificity and potency (29–32). Venetoclax (ABT-199; GDC-0199/VENCLEXTA) is an FDA approved BCL-2 inhibitor for use in (relapsed) chronic lymphocytic leukemia (30,31). Venetoclax has also shown clinical efficacies against acute myeloid leukemia due to its activity against leukemic stem cells (32), and has recently been used in clinical trials of breast cancer treatment (33,34).

Given our strong preclinical data showing that BCL-2 is induced by ARPIs to causally drive castration resistance, that BCL-2 is upregulated in diverse and heterogeneous CRPC subtypes and models, and that BCL-2 inhibitor ABT-199, when used alone or in combination with enzalutamide, displays strong CRPC-inhibitory effects (11,35), we propose that simultaneously targeting both AR^+^ and AR^−/lo^ CRPC cell populations from the outset, rather than sequentially, may offer therapeutic advantages and help delay the development of resistance to second-generation ARPIs. Here we present the results from a prospective phase Ib clinical trial, I-63418 (NCT03751436), which demonstrate the feasibility of this novel therapeutic combination, provide important insights into the pharmacokinetic (PK) interactions between venetoclax and enzalutamide and potential clinical benefits, and present proof-of-principle findings that venetoclax promotes rapid BCL-2β generation and subsequent BCL-2 protein degradation.

## Materials and Methods

### Study design and objectives

I-63418 was a phase Ib open label single-arm, single-center study of enzalutamide with venetoclax (ABT-199) in patients with mCRPC (NCT03751436). The study protocol was approved by the institutional review board (IRB) and ethics committee prior to initiation and conducted in accordance with the declaration of Helsinki, and patients were provided with written informed consent before entering the study. A total of 10 patients were enrolled from Aug. 2019 to Sept. 2022 per a standard 3+3 design, starting with a standard dose of enzalutamide (160 mg/d) and 3 dose levels (DL) of venetoclax at 400 mg/d (DL1), 600 mg/d (DL2) and 800 mg/d (DL3) (Supplementary Fig. S1).

The primary objectives of this phase Ib study were to: 1) characterize the safety and tolerability profile and 2) determine the dose-limiting toxicity (DLT), maximum tolerated dose (MTD) and recommended phase II dose (RP2D) of the enzalutamide and venetoclax combination in patients with mCRPC. We also analyzed the pharmacokinetic (PK) profiles of enzalutamide and venetoclax when given in combination to confirm whether drug levels were in the therapeutic range. We additionally performed pharmacodynamic (PD) studies using patients’ and healthy donor peripheral blood mononuclear cells (PBMCs) to determine how venetoclax (ABT199) treatment impacted the target (BCL-2) expression.

### Patient eligibility

Patients aged 18 years or older with histologically confirmed prostate adenocarcinoma were eligible for enrollment. All subjects had mCRPC exhibiting progressive disease, as evidenced by rising prostate-specific antigen (PSA) levels and/or radiographic progression. Eligible patients had adequate organ function and an Eastern Cooperative Oncology Group (ECOG) Performance Status of 0 or 1. Prior cytotoxic chemotherapy in the metastatic or castration-refractory setting was permitted. However, key exclusion criteria included untreated or active central nervous system metastases, current active hematological malignancies, previous treatment with BCL-2 inhibitors, anticancer therapy within 21 days before the first cycle, radiotherapy within 21 days or to the target lesion sites.

### Study endpoints and assessments

Enzalutamide was administered at a standard dose of 160 mg (40 mg x 4 pills) orally once daily. After a minimum of 4 weeks of exposure to enzalutamide (lead-in time), venetoclax was administered at a starting dose of 400 mg and escalated from 400 mg to 600 mg, and then from 600 mg to 800 mg, in cohorts of 3 patients (Supplementary Fig. S1). The primary endpoint was to determine the MTD for venetoclax based on the rate of DLTs. Adverse events (AEs) were categorized and graded according to NCI CTCAE version 5.0. The RP2D was to be selected based on overall tolerability of the regimen, and PK analysis of plasma enzalutamide and venetoclax concentrations at specified time points was performed.

Patients had a CT scan of the chest, abdomen, and pelvis, and a bone scan at baseline. Tumor biopsy was performed as dictated by standard of care in the presence of soft tissue or visceral metastatic disease; however, it was not mandatory for enrollment. Response and progression assessments were made using a combination of the international criteria proposed by the Response Evaluation Criteria in Solid Tumors version 1.1 (RECIST 1.1) and the guidelines for PCa endpoints developed by the Prostate Cancer Clinical Trials Working Group (PCWG3). The same radiographic procedure used to assess disease sites at screening was used throughout the study. PSA measurement and radiographic assessment were done at 12-week intervals. All enrolled mCRPC patients had tumor assessments conducted regardless of dose delays or early discontinuation, pre-defined study end (at least 1 year after they have discontinued study treatment), or until death, whichever occurred first.

### PK studies

We characterized PK profiles of enzalutamide and venetoclax when given in combination to patients with mCRPC. Venetoclax treatment began on day 1, after patients had reached steady-state enzalutamide levels following 4 weeks of lead-in treatment (Supplementary Fig. S1). Blood samples were collected in EDTA tubes prior to and at 1, 2, 4, 6, 8, and 24 h post venetoclax on days 1 and 29. Additional samples were collected prior to dosing on days 8, 15, 21, and 57 (Supplementary Fig. S1). Samples were centrifuged at approximately 1,000 x g, and plasma was stored at −70°C or colder until analysis.

All samples were analyzed for venetoclax concentrations with an LC-MS/MS assay validated to FDA guidance over the range of 5-5,000 ng/mL. A volume of 50 μL of the standard, QC, or sample plasma was pipetted into a microfuge tube and 10 μL of internal standard (IS; 500 ng/mL ^2^H_7_-venetoclax) was added to each. Next, 200 μL of acetonitrile was added followed by vortexing for 30s. Samples were centrifuged at 17,200 × g at room temperature for 10 min. Supernatants were transferred to autosampler vials, followed by injection of 5 μL into the LC-MS/MS system. The LC system consisted of an Exion autosampler with binary pump (Sciex, Framingham, MA, USA), Synergi Polar-RP C18 (4 µm, 2.1 x 50 mm) column kept at 30°C. The autosampler temperature was also kept at 10°C. Mobile phase solvent A consisted of 0.1% formic acid in water and mobile phase solvent B consisted of 0.1% formic acid in acetonitrile, mixed at 60% B. A. flow rate of 0.4 mL/min between 0-3 min and 1 mL/min from 3-5 min. Retention time of venetoclax was 1.7 min. MS detection was carried out using an ABI SCIEX (Concord, ON, Canada) 4500 tandem mass spectrometer with electrospray ionization in positive multiple reaction monitoring (MRM) mode, monitoring *m/z* transitions: 868.40>836.3 for venetoclax and 875.4>643.3 for venetoclax IS. QC based accuracies (N=6, each of 3 days) were 100.9-106.2%. The intra- and inter-assay precisions were <10.6% and <2.4%. Incurred sample reanalysis of 27 samples yielded the following results (%samples with a difference larger than 20% / median difference / median absolute difference): 0% / −3.3% / 3.4%.

All trough samples were analyzed for enzalutamide and metabolite concentrations with an LC-MS/MS assay validated to FDA guidance (36). Incurred sample reanalysis of 19 samples yielded the following results (%samples with a difference larger than 20% / median difference / median absolute difference) 5% / 4.3% / 7.1% for enzalutamide and 0% / 0.9% / 3.6% for N-desmethyl enzalutamide.

Plasma PK parameters were derived from the data by non-compartmental methods with WinNonlin (Certara, Princeton, NJ). Statistical analyses for PK parameter values were performed using SPSS 27.0 for Windows (SPSS Inc., Chicago, IL). Within patient variability in C_min_ was calculated from the day 8 through day 57 data across doses by calculating 100 × the square root of the mean square within groups obtained after ANOVA of log-transformed values of these parameters with subject as a factor.

### PBMC isolation

PBMCs from the trial patients or healthy donors were isolated from whole blood using Lymphoprep™ (STEMCELL Technologies, Cat. #07801) following the manufacturer’s protocol. Whole blood was diluted 1:1 with room temperature phosphate-buffered saline (PBS) containing 2% fetal bovine serum (FBS) and carefully layered over an equal volume of Lymphoprep™ in a 50 mL conical tube. Samples were centrifuged at 800 × g for 20–30 minutes at room temperature with the brake off. The PBMC layer formed at the interface between the plasma and Lymphoprep™ was gently collected and transferred to a new tube. Cells were washed once with PBS containing 2% FBS by centrifugation at 300 × g for 10 minutes, and the final pellet was resuspended in DMEM media for downstream culture.

### Automated Western Blot (WES) analysis of BCL-2 protein in patient PBMCs

The BCL-2 protein levels in patient PBMCs were analyzed using PBMC lysate in the automated quantitative WES^TM^ system (https://www.bio-techne.com/brands/proteinsimple), as we previously described (37). Briefly, the Simple Western WES^TM^ immunoassays take place in capillaries and separate proteins by size as they migrate through stacking and separation matrix. The separated proteins are immobilized to the capillary wall via a proprietary, photoactivated capture chemistry. The target protein was identified using a primary antibody and immunoprobed using an HRP-conjugated secondary antibody and chemiluminescent substrate. The resulting chemiluminescent signal is displayed as traditional virtual blot-like image and electropherogram. Quantitative results such as M.W, signal intensity (area), % area, and signal-to-noise for each immunodetected protein are presented in the results table automatically.

For our studies herein, proteins were extracted from patient PBMCs using 0.1× WES Sample Buffer (ProteinSimple; cat#042-195) following the manufacturer’s instructions to preserve protein integrity and compatibility with the WES system. Protein concentrations were measured using the BCA Protein Assay Kit (Thermo Fisher Scientific, cat#23225), and equal amounts of total protein (0.375 µg per sample) were mixed with 5× fluorescent master mix, denatured at 95°C for 5 minutes, and 3 μl of each sample was loaded into individual capillaries of the WES system. The instrument performed automated size-based protein separation, blocking, incubation with primary and secondary antibodies, and chemiluminescent detection. BCL-2 was detected using a mouse mAb (clone 124, Cell Signaling Technology, cat#15071S) diluted 1:100 in Antibody Diluent (ProteinSimple) while β-actin served as the internal loading control and was detected using a rabbit pAb (Novus Biologicals, cat# NB600-503) diluted 1:25.

### Treatment of healthy donor PBMCs *ex vivo* with ABT-199

ABT-199 (S8048, Selleck Chemicals) was dissolved in DMSO. Healthy donor PBMCs were seeded at a density of 2 × 10⁶ cells per 60 mm dish and treated with 1 μM of ABT-199 for various time intervals (i.e., 2, 4, 8, and 24 h). At the end of each time point, PBMCs were harvested for protein and RNA extraction (below). All treatments were performed in 3-6 biological replicates and control samples were treated with vehicle (DMSO) only. In all experiments, the vehicle concentration did not exceed 0.2%.

### Analysis of BCL-2 α/β proteins by Western blotting

PBMC samples from above were harvested using 0.1% trypsin-EDTA solution (Gibco) and centrifuged (1,000 rcf, 4 min, 4°C), washed with DPBS (Gibco), centrifuged again (1,000 rcf, 4 min, 4°C), and the pellet was lysed in denaturing buffer containing Halt protease inhibitor cocktail (Thermo Fisher Scientific; Cat# SD-001/SN-002) for 15 min on ice. After centrifugation (15,000 rcf, 15 min, 4°C), an aliquot of the supernatant was taken to determine the protein concentration using a BCA Protein Assay Kit (Thermo Fisher Scientific). Protein samples were mixed in NuPAGE LDS buffer and 1x reducing agent (Invitrogen) and denatured at 95°C for 5 min. Proteins separated by sodium dodecyl sulfate–polyacrylamide gel electrophoresis (SDS-PAGE) were transferred to a nitrocellulose membrane at 110 V for 110 min in a wet tank transfer system (Surelock system, Invitrogen). Then, the membranes were incubated in a 5% solution of nonfat milk powder in TBST buffer (20 mM Tris, 150 mM NaCl, 0.025% Tween 20, pH 7.4) for 45 min at room temperature to block nonspecific protein binding. After blocking, the membranes were washed in TBST (4 × 5 min) and incubated with primary antibody at 4°C for ∼16 h. The membranes were washed in TBST (4 × 5 min), and secondary antibody was added in 2.5% nonfat milk powder solution and incubated for an hour at room temperature. After washing (4 × 5 min) in TBST, membranes were incubated in BrightStar™ (Alkali scientific) and Chemiluminescence detection and image analysis were performed using the ChemiDoc XRS^+^ System (Bio-Rad). The following primary antibodies were used at 1:1000 dilution: mouse mAb against BCL-2 (Cell Signaling Technology, cat# 15071), rabbit pAb against BCL-2β (Bioss Antibodies, Woburn, MA, USA, cat# bs-15534R), rabbit pAb to cleaved CASP3 (Cell Signaling Technology, cat#9661), rabbit mAb to GAPDH (Cell Signaling Technology, cat#2118, clone 14C10), and rabbit mAb to β-actin (clone 13E5, HRP Conjugate; cat#5125).

### qPCR analysis of BCL-2 α/β isoform mRNAs

We developed a qPCR-based strategy to quantitatively analyze the *BCL-2α/β* mRNA levels in control and ABT-199 treated PBMCs (above). Briefly, total RNA was extracted using the PicoPure™ RNA Isolation Kit (Thermo Fisher Scientific, cat# KIT0204) with on-column DNase treatment. cDNA was synthesized from 750 ng RNA using SuperScript™ IV VILO™ Master Mix (Thermo Fisher Scientific, cat# 11756050). Quantitative PCR was performed using TaqMan Gene Expression Assays (Bio-Rad) with a FAM-conjugated custom probe specific for the BCL-2β isoform (5’-CTGAGGCCACAGGTCCGAG-3’; Bio-Rad, part# 10031261). The forward and reverse primers were 5’-GGTGCACTTGGTGATGTGA-3’ and 5’-CTCCACAGCCTCCCATTG-3’, respectively, which quantifies a 127bp amplicon (5’-TAGGTGCACTTGGTGATGTGAGTCTGGGCTGAGGCCACAGGTCCGAGATGCGGGGGTTGGAGTGCGGGTGGGCTCCTGGGGCAATGGGAGGCTGTGGAGCCGGCGAAATAAAATCAGAGTTGTTGCT-3’).

β-Actin (ACTB) was used as the internal control (Applied Biosystems, cat# 4326315E). Reactions were run on the QuantStudio™ 6 Flex Real-Time PCR System (Applied Biosystems) under standard cycling conditions. Relative expression was calculated using the 2^–ΔΔCt method normalized to the 0-hour (untreated) control. All conditions were tested in technical triplicates from 3-6 independent experiments.

### Statistical analyses

The intention-to-treat population comprised of all enrolled patients regardless of study treatment administration: the safety-evaluable population included all patients who received any study drug. The primary analysis was conducted 6 months after the last patient was enrolled in the study. Safety analysis was descriptive in the safety evaluable population, and pre-defined methodology was used for toxicity reporting. DLTs are reported as descriptive statistics. All adverse events were summarized by the treatment arm, with number of patients and percentages. The efficacy of the combination was evaluated on the PCWG3 criteria for response to treatment, and objective tumor response was tabulated overall (and by dose level if appropriate). Only participants who had completed at least 2 cycles were evaluated for response. Kaplan-Meier survival analysis was conducted to estimate median progression-free survival (PFS), overall survival (OS), and time to next systemic therapy (TNST).

Statistical analyses for PK studies were described above. For statistical analysis of the ABT-199 effects on BCL-2 protein expression in PBMCs, between 3 to 6 independent biological replicates were used for each experimental time point, and BCL-2 protein levels across groups were compared using the Kruskal-Wallis test. Fold change ratios in BCL-2β/α were calculated, and statistical significance was assessed using a one-sample two-tailed *t-*test. For qPCR, we used one-sample two-tailed *t*-tests to compare each group mean to a reference value of 1, which was assigned to the 0 h sample. Statistical significance was defined as *p* < 0.05. Asterisks “*”, “**”, and “***” denote *p* < 0.05, *p* < 0.01, and *p* < 0.001, respectively. All statistical analyses were conducted using GraphPad prism.

### Data Availability

Access to individual patient level clinical data as well as all correlative and biochemistry data can be made by contacting the corresponding authors.

## Results

### Patient demographics

Ten patients were enrolled in study I-63418 between 2019-2022 with median age at 71.3 years (range: 59.1-76.6) (**Table 1**). There were 3, 3, and 4 patients enrolled for venetoclax DL1 (400 mg), DL2 (600 mg) and DL3 (800 mg), respectively. All patients had received prior chemotherapy. Nine patients had progressed on prior ARPI treatment, and seven patients had prior exposure to radiation. While most patients completed the study, one patient at DL3 had to be replaced as he chose to come off study prior to the 30-day endpoint.

**Table 1.** Baseline patient characteristics for I-63418 (NCT03751436)

| **Variable** |  | **DL-1 %**<br>**(n)** | **DL-2 %**<br>**(n)** | **DL-3 %**<br>**(n)** | **Overall %**<br>**(n)** |
| --- | --- | --- | --- | --- | --- |
| Number of Patients |  | 30.0 (3) | 30.0 (3) | 40.0 (4) | 100 (10) |
| Median Age |  | 73.2 | 69.4 | 69.1 | 71.3 |
| Caucasian |  | 100 (3) | 100 (3) | 100 (4) | 100 (10) |
| Adenocarcinoma |  | 100 (3) | 100 (3) | 100 (4) | 100 (10) |
| ECOG | 0 | 33.3 (1) | 33.3 (1) | 75.0 (3) | 50.0 (5) |
|  | 1 | 66.7 (2) | 66.7 (2) | 25.0 (1) | 50.0 (5) |
| Prior Surgery |  | 33.3 (1) | 0 | 75.0 (3) | 40.0 (4) |
| Prior Chemo |  | 100.0 (3) | 100.0 (3) | 100.0 (4) | 100 (10) |
| Prior Radiation |  | 100.0 (3) | 100.0 (3) | 25.0(1) | 70.0 (7) |
| Prior AR targeted therapy |  | 66.6 (2) | 100.0 (3) | 100.0 (4) | 90.0 (9) |

### Combination of venetoclax with enzalutamide was well tolerated

No DLTs were observed at DL1 - DL3; however, grade 3 fatigue and thromboctytopenia was observed in one patient, respectively, at DL3 (**Table 2**). In general, the combination was very well tolerated with most adverse events being low grade (β2). The most frequent treatment related adverse event (TRAE) was fatigue, affecting 90% of patients. Overall, 50% of the patients experienced grade 2 TRAEs while 30% of the patients experienced grade 1 TRAE (**Table 2**).

**Table 2.** Drug-related adverse events observed in patients from I-63418 (NCT03751436)

| **Adverse events** | **Grade 1 %(n)** | **Grade 2 %(n)** | **Grade 3 %(n)** | **All % (n)** |
| --- | --- | --- | --- | --- |
| Gastroesophageal reflux disease | 20 (2) | 0 | 0 | 20 (2) |
| Nausea | 30(3) | 0 | 0 | 30(3) |
| Anorexia | 10(1) | 10(1) | 0 | 20(2) |
| Hyperglycemia | 10(1) | 0 | 0 | 10(1) |
| Dizziness | 10(1) | 0 | 0 | 10(1) |
| Dysgeusia | 10(1) | 0 | 0 | 10(1) |
| Headache | 20(2) | 0 | 0 | 20(2) |
| Fatigue | 40(4) | 40(4) | 10(1) | 90(9) |
| Thrombocytopenia | 0 | 0 | 10(1) | 10(1) |
| Pain | 10(1) | 0 | 0 | 10(1) |
| Arthralgia | 0 | 10(1) | 0 | 10(1) |
| Hot Flashes | 20(2) | 0 | 0 | 20(2) |
| Insomnia | 10(1) | 0 | 0 | 10(1) |

### Venetoclax exposure did not increase upon dose escalation

We conducted PK analysis on samples from each dose level to better understand the relationship and potential drug-drug interactions between venetoclax and enzalutamide. As venetoclax dose increased from 400 to 800 mg, overall patient exposure (AUC_0-last_) did not proportionally increase with dose (**Fig. 1A-C**; **Table 3**). The ratio of day 29 AUC_0-24_ over day 1 AUC_0-inf_ was 0.77 (2.90) (geometric mean and geometric standard deviation; corresponding to a 190%CV). Although there was large variability, the C_min_ concentrations did not appear to vary over time with continued dosing, nor did C_min_ levels appear to increase much with increasing dose (**Fig. 1D-F**; **Table 3**). Intrapatient variability in C_min_ was calculated at 51%CV from day 8 through 57, which was less than the interpatient variability approximated of 117%CV (based on geometric standard deviation: (2.17-1)*100=117%) (**Table 3**).

**Fig. 1.**
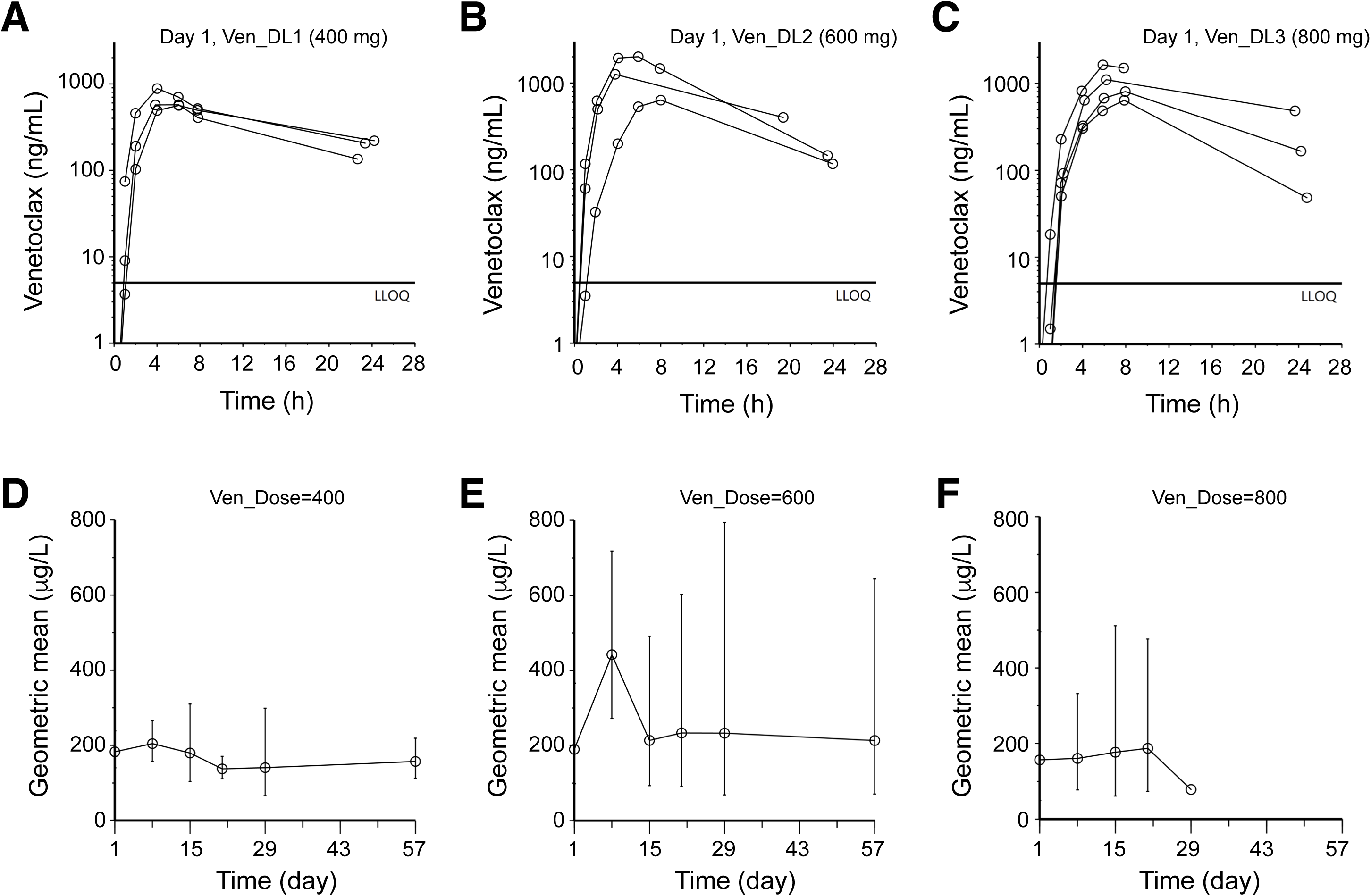
PK studies of venetoclax. Shown in (**A-C**) are individual PK profiles of venetoclax after doses of 400 mg (A), 600 mg (B) or 800 mg (C) on day 1. Shown on (**D-F**) are trough venetoclax concentrations (geometric mean ± geometric standard deviation) over the period of study with venetoclax dosing starting 24 h before the day 1 data point.

**Table 3.** Venetoclax plasma PK parameters (geometric mean (geometric standard deviation)).

|  | D1 |  |  |  |  |  |  | D15 | D29 |  |  |
| --- | --- | --- | --- | --- | --- | --- | --- | --- | --- | --- | --- |
| Dose<br>(mg) | C<sub>max</sub><br>(µg/L) | C<sub>max</sub>/Dose<br>(µg/L/mg) | T<sub>max</sub><br>(h) | t<sub>1/2</sub><br>(h) | AUC<sub>0-last</sub><br>(mg/L•h) | AUC<sub>0-inf</sub><sup>a</sup><br>(mg/L•h) | Cl/F<br>(L/h) | C<sub>min</sub><br>(µg/L) | C<sub>max</sub><sup>b</sup><br>(µg/L) | AUC<sub>0-24</sub><sup>b</sup><br>(mg/L•h) | Ratio<sup>c</sup><br>D29/D1 |
| 400<br>(N=3) | 658<br>(1.29) | 1.64 (1.29) | 5.2<br>(1.3) | 11<br>(1.2) | 8.02 (1.21) | 11.1 (1.27) | 36.2<br>(1.27) | 180<br>(1.72) | 968<br>(1.67) | 10.5 (1.65) | 0.95<br>(1.99) |
| 600<br>(N=3) | 1172<br>(1.79) | 1.95 (1.79) | 5.6<br>(1.5) | 6.6<br>(1.4) | 12.2 (1.65) | 14.6 (1.65) | 41.2<br>(1.65) | 214<br>(2.30) | 554<br>(4.89) | 9.11 (4.1) | 0.62<br>(5.32) |
| 800<br>(N=4) | 979<br>(1.50) | 1.22 (1.50) | 6.9<br>(1.2) | 9.4<br>(1.8) | 8.76 (1.54) | 16.6 (2.31) | 48.1<br>(2.31) | 177<br>(2.89) | 1000 (-) | 8.87 | 0.79 |
| Total<br>(N=10) | 917<br>(1.57) | 1.54 (1.54) | 6.0<br>(1.3) | 8.9<br>(1.6) | - | - | 42.1<br>(1.76) | 188<br>(2.17) | 765<br>(2.74) |  | 0.77<br>(2.90) |

<sup>a</sup>AUC<sub>0-inf</sub> extrapolated beyond the last time point sampled was geometric mean 20% (range 4.9-84%).
<sup>b</sup>N=3, 3, and 1 respectively for the 400, 600, and 800 mg dose.
<sup>c</sup>Ratio of day 29 AUC<sub>0-24</sub> over day 1 AUC<sub>0-inf</sub>.

Upon initiation of venetoclax treatment, enzalutamide and N-desmethyl enzalutamide trough appeared to increase slightly, but only the increase in enzalutamide trough concentrations reached statistical significance (Supplementary Fig. S2; Supplementary Table S1). Intrapatient variability in C_min_ was calculated as 7.0%CV from day 1 through 57 for enzalutamide and 10%CV for N-desmethyl enzalutamide, suggesting that their levels were stable (Supplementary Table S1).

In aggregate, the PK studies suggest minimal impact of venetoclax on enzalutamide metabolism but sub-therapeutic plasma levels of venetoclax in mCRPC patients treated with the combination.

### Enzalutamide combined with venetoclax produced modest clinical response

Median treatment duration on this trial was 11.4 weeks (**Table 4**). A best response of stable disease was observed in four patients (40%) while six patients had progressive disease (60%) (**Table 4**). Interestingly, two out of three patients at DL2 showed prostate-specific antigen (PSA; Supplementary Fig. S3A-B) and radiographic (data not shown) responses. Estimated median progression-free survival (PFS) and overall survival (OS) were 2.6 months and 18.8 months, respectively (**Table 4**).

**Table 4.**
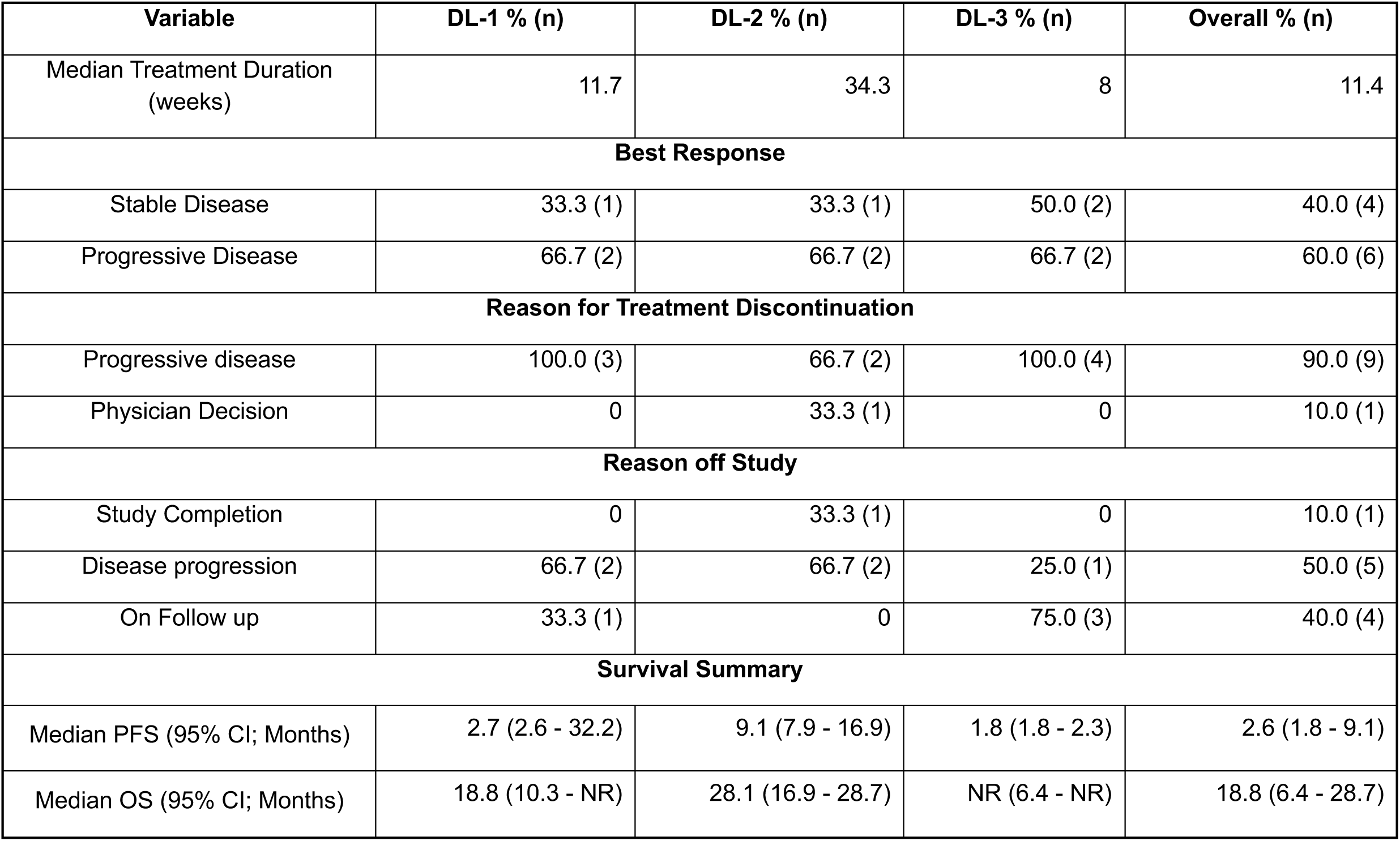
Measures of efficacy.

| Variable | DL-1 % (n) | DL-2 % (n) | DL-3 % (n) | Overall % (n) |
| --- | --- | --- | --- | --- |
| Median Treatment Duration (weeks) | 11.7 | 34.3 | 8 | 11.4 |
| **Best Response** |  |  |  |  |
| Stable Disease | 33.3 (1) | 33.3 (1) | 50.0 (2) | 40.0 (4) |
| Progressive Disease | 66.7 (2) | 66.7 (2) | 66.7 (2) | 60.0 (6) |
| **Reason for Treatment Discontinuation** |  |  |  |  |
| Progressive disease | 100.0 (3) | 66.7 (2) | 100.0 (4) | 90.0 (9) |
| Physician Decision | 0 | 33.3 (1) | 0 | 10.0 (1) |
| **Reason off Study** |  |  |  |  |
| Study Completion | 0 | 33.3 (1) | 0 | 10.0 (1) |
| Disease progression | 66.7 (2) | 66.7 (2) | 25.0 (1) | 50.0 (5) |
| On Follow up | 33.3 (1) | 0 | 75.0 (3) | 40.0 (4) |
| **Survival Summary** |  |  |  |  |
| Median PFS (95% CI; Months) | 2.7 (2.6 - 32.2) | 9.1 (7.9 - 16.9) | 1.8 (1.8 - 2.3) | 2.6 (1.8 - 9.1) |
| Median OS (95% CI; Months) | 18.8 (10.3 - NR) | 28.1 (16.9 - 28.7) | NR (6.4 - NR) | 18.8 (6.4 - 28.7) |

Overall, one patient (1/10; 10%) achieved PSA50 (patient 01-03; Supplementary Fig. S3B). However, four patients showed both PSA and radiographic responses, including one patient at DL-1 (33%; patient 01-03), two patients at DL-2 (67%; patients 02-01 and 02-07) and one patient at DL-3 (25%; patient 03-05), respectively (Supplementary Fig. S3B; data not shown). The DL-1 cohort had a mean PSA of 28.1 ng/mL at baseline, which increased to 197.5 ng/mL by the end of treatment. The DL-2 cohort had a mean PSA of 21.8 ng/mL at baseline, which only increased slightly to 29.6 ng/mL by the end of treatment. Lastly, the DL-3 cohort had a mean PSA of 92.1 ng/mL at baseline, which increased to 166.0 ng/mL by the end of treatment (Supplementary Fig. S3A). Although these results remain inconclusive due to the nature of this trial design, some patient response to combination treatment was observed.

### Venetoclax (ABT-199) promoted BCL-2β generation and BCL-2 degradation in PBMCs

We performed correlative and biochemistry studies to gain insight on how venetoclax (ABT-199) might have affected the expression of its target, BCL-2 (**Fig. 2**). BCL-2 has two major mRNA splicing variants, i.e., *BCL-2α* (NM_000633.3; encoding the commonly known 26-kDa BCL-2 protein) and *BCL-2β* (NM_000657.3) (**Fig. 2A**). BCL-2β protein lacks the transmembrane (TM) domain (**Fig. 2A**) and has much reduced anti-apoptotic functions, and BCL-2β/BCL-2α ratio has been proposed to be a ‘rheostat’ for apoptosis initiation (38–40). Automated quantitative analysis using the WES system (see Methods; 37) in patients’ PBMCs from two patients at DL2 (i.e., 02-01-RP and 02-07-RP) and one patient at DL1 (01-03-RP) revealed that venetoclax promoted the generation of BCL-2β with concordant reduction in BCL-2α protein during early cycles of treatment (**Fig. 2B**). In late cycles and towards the end of treatment (EoT), both BCL-2α and BCL-2β were markedly reduced (**Fig. 2B**) accompanied, frequently, by the appearance of lower M.W degradation products (**Fig. 2B**; asterisks).

**Fig 2.**
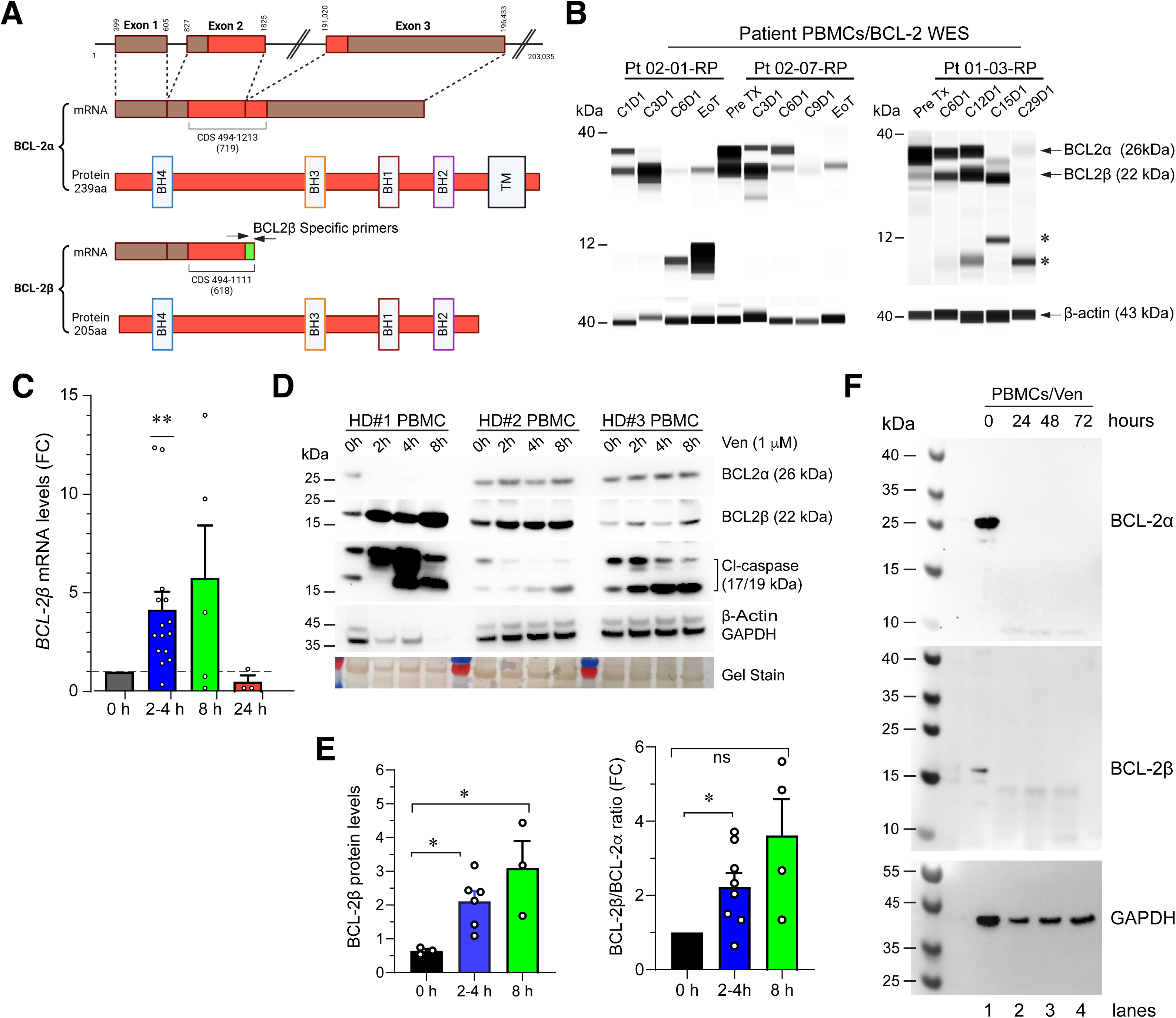
Venetoclax promoted BCL-2β generation and BCL-2α/BCL-2β protein degradation in both patients’ and healthy donor (HD) PBMCs. **(A)** Schematic of the *BCL-2* genomic structure with 3 exons (top) and the mRNA and protein structures of the two major BCL-2 splice isoforms, i.e., BCL-2α and BCL-2β. Although both BCL-2α and BCL-2β have 4 BH (BCL-2 homology) domains, BCL-2α includes a C-terminal transmembrane (TM) domain needed for its anti-apoptotic function at the mitochondria whereas BCL-2β lacks this domain making it a shorter, mostly cytosolic protein lacking anti-apoptotic activity. To detect *BCL-2β* mRNA specifically, we designed primers targeting the unique exon-exon junction only present in the *BCL-2β* transcript (arrows). **(B)** Automated WES analysis of venetoclax-induced BCL-2β generation and BCL-2 degradation in PBMCs from 3 trial patients (Pt), who also showed PSA responses (see Supplementary Fig. S3B). PBMCs were isolated from whole blood collected at pretreatment (Pre TX), D1 of the indicated cycles (C) or end of treatment (EoT). Note patient-specific changes in BCL-2α/β protein levels. **(C)** In healthy donor (HD) PBMCs, Venetoclax (ABT-199, 1 µM) caused time-dependent changes in *BCL-2β* mRNA levels. Briefly, the *BCL-2β* mRNA levels increased significantly at 2–4 h post-treatment (**p < 0.01) with a trend toward increase at 8 h but reduced at 24 h post treatment. Each dot represents an independent HD PBMC treated with ABT-199 (i.e., biological replicates) and the results were presented as fold change (FC) over the untreated samples (0 h). **(D)** Venetoclax (Ven) induced dynamic and donor-dependent BCL-2β and BCL-2α protein changes in HD PBMCs. In HD#1 PBMCs, venetoclax caused rapid (i.e., within 2 h) and persistent (up to 8 h) induction of BCL-2β with concomitant loss of BCL-2α leading to significant apoptosis (i.e., elevated Cl-caspase-3) such that even the loading control proteins β-actin and GAPDH were decreased/lost by 8 h. In HD#2 PBMCs, venetoclax increased BCL-2β without significant changes in BCL-2α and increased Cl-caspase within 4-8 h. In HD#3 PBMCs, venetoclax caused BCL-2β upregulation (without significant changes in BCL-2α) and increased apoptosis at around 2 h post treatment. Shown at the bottom is an image of the gel stained by Swift stain (as another loading control). Note that low levels of Cl-caspase-3 were observed in untreated (i.e., 0 h) PBMCs due to stress from the isolation process. **(E)** Quantification of BCL-2β protein levels (left) and BCL-2β/BCL-2α ratio as fold changes (FC; right) in HD PBMCs treated with venetoclax (i.e., ABT-199). Results represent the aggregated data from independent experiments (n=3-7) exemplified in D. **(F)** Western blotting showing that both BCL-2α and BCL-2β proteins were lost in HD PBMCs at β24 h post venetoclax (Ven; 1 μM) treatment.

We further studied the venetoclax effects on BCL-2α/BCL-2β in PBMCs freshly purified from healthy donors (HD) by employing a custom-designed *BCL-2β* specific (**Fig. 2A**) qPCR assay (**Fig. 2C**) and using a BCL-2β specific antibody (**Fig. 2D-F**). Interestingly, venetoclax (ABT-199) at 1 μM rapidly (i.e., within 2-4 h) induced *BCL-2β* mRNA levels for up to 8 h (**Fig. 2C**; note *BCL-2β* mRNA levels at 8 h were apparently, although not statistically significantly, increased). Western blotting showed that in all 3 HD PBMCs, venetoclax upregulated BCL-2β protein levels and increased the BCL-2β/BCL-2α ratios as early as 2 h post treatment and the effect lasted for up to 8 h (**Fig. 2D-E**). Intriguingly, in HD#1 PBMCs, venetoclax caused rapid and huge increases in BCL-2β with concomitant loss of BCL-2α whereas in HD#2 and HD#3 PBMCs, venetoclax induced more modest BCL-2β without major changes in BCL-2α levels (**Fig. 2D**). Nevertheless, in PBMCs from all 3 HDs, venetoclax caused apoptosis as evidenced by increased activated (cleaved) caspase-3 (Cl-caspase; **Fig. 2D**). At ≥24 h post venetoclax treatment, both *BCL-2β* mRNA (**Fig. 2C**) and BCL-2α/BCL-2β proteins (**Fig. 2F**) became undetectable.

## Discussion

The current phase Ib combination trial with enzalutamide and venetoclax in patients with mCRPC was motivated by our preclinical data that implicates BCL-2 as a strong driver of castration resistance (11,35), availability of potent and selective BCL-2 inhibitor venetoclax, clinical approval of venetoclax in hematological malignancies (30–32), and emerging clinical trials of venetoclax in solid tumors such as breast cancer (33,34). The combination of enzalutamide and venetoclax demonstrated a manageable safety profile with the most common TRAEs being fatigue (90%), nausea (30%), and hot flashes (20%), which were predominantly low-grade (≤2). Only one patient experienced grade 3 fatigue at the highest DL3 of 800 mg venetoclax. This favorable safety profile is encouraging, as it suggests the potential for combining ARPIs with venetoclax without excessive toxicity.

While the study was primarily focused on safety and tolerability, preliminary efficacy data showed promising signals. Four patients (40%) achieved stable disease as their best response, with two out of three patients in the 600 mg venetoclax DL2 (i.e., patients 02-01 and 02-07) experiencing stable disease with PSA and radiographic response. This observation aligns with the proposed mechanism of action, wherein enzalutamide targets the AR^+^ PCa cells and venetoclax targets primarily AR^−/lo^ cell population, potentially delaying or preventing resistance to second-generation ARPIs. Although promising, a follow up phase II study is required to further test this hypothesis. The median PFS of 2.6 months and median OS of 18.8 months observed in this study are consistent with the expected outcomes in heavily pretreated mCRPC patients. Admittedly, one patient at venetoclax DL1 (i.e., 01-03) had limited exposure to AR-targeted therapy prior to study entry, had prolonged stable disease, and may have favorably skewed the PFS analysis.

PK analysis revealed that venetoclax clearance was estimated at approximately 42 L/h or 1008 L/day, which is over twice the value of 413 L/day previously reported (41). Venetoclax half-life was found to be approximately 9 h, also far shorter than the 14-26 h values previously reported (32). Indeed, previous C_max_ and C_min_ values of 3740 and 1430 µg/L, as well as normalized C_max_ values of 4-6 µg/L associated with an 800 mg dose, are all higher than results observed in the current trial (32). Enzalutamide is a strong CYP3A4 inducer, which increased the oral clearance of midazolam, a sensitive CYP3A4 substrate, by 7-fold (42). CYP3A4 plays a major role in venetoclax clearance, and rifampin, the typical CYP3A4 inducer, increased venetoclax clearance approximately 6-fold (43). Data presented here suggest that enzalutamide co-administration leads to a 2-3-fold reduction in venetoclax exposure and sub-therapeutic venetoclax levels in mCRPC patients. A venetoclax exposure-response relationship has been established in CLL with a doubling of dose extending PFS by 6 months and increased average venetoclax exposure resulting in increased likelihood of an objective response (44,45). Furthermore, in multiple myeloma the AUC_0-24_ was positively correlated with the probability of response (46). The exposure metrics observed in the current trial were in the lower ranges of those described in previous exposure-response relationships. The less than proportional increase in exposure with dose observed in our study would appear to be more severe than the 12% less than expected exposure when doubling dose from 400 to 800 mg as characterized in a population PK report (41). This observation implies that *active levels of venetoclax may not be achieved even when further increasing the venetoclax dose in combination with enzalutamide*.

On the other hand, enzalutamide pharmacokinetics observed in the current trial was in line with previous reports for both enzalutamide (11.7 vs 11.4 µg/L) and N-desmethyl enzalutamide (11.0 vs 13 µg/L) (47). The observed increase in enzalutamide of approximately 9% upon initiation of venetoclax treatment may be due to either competitive inhibition of CYP3A4 or a local effect on the absorption process in the gut. While statistically significant, this is not clinically relevant given the small effect size observed and the pharmacological contributions of the N-desmethyl enzalutamide active metabolite.

Despite the pharmacokinetic limitations, our study observed promising signals of clinical activity, with 40% of patients achieving stable disease as their best response. Significantly, biochemical and molecular studies reveal that even at sub-therapeutic levels, venetoclax ‘hit’ its target, BCL-2. In both patients’ and healthy donor PBMCs, venetoclax treatment led to rapid increase of the BCL-2β isoform followed, later, by degradation (loss) of both BCL-2α and BCL-2β. Lacking the critical TM domain, BCL-2β has lost anti-apoptotic activity and BCL-2β/BCL-2α ratio may set the threshold for initiating the intrinsic cell death pathway (38–40). In support, our results show, in venetoclax-treated PBMCs, increased caspase activity with increasing BCL-2β expression and loss of BCL-2 proteins. These effects may underlie the promising clinical response in several patients, specifically, patients 01-03, 02-01 and 02-07, all of whom also exhibited PSA responses. In a companion paper, we represent evidence that enzalutamide and venetoclax combination also reduced multiple molecular targets as well as circulating tumor cells in these ‘responsive’ patients (35).

In summary, this phase Ib study provides valuable insights into the potential of combining enzalutamide and venetoclax in treating patients with mCRPC while also highlighting the challenges and opportunities for future research. The favorable safety profile and preliminary efficacy signals warrant further investigations of combining ARPIs and venetoclax. While our study demonstrated the challenges of achieving therapeutic venetoclax levels when combined with enzalutamide, the importance of targeting the BCL-2 pathway in mCRPC remains evident. Given the untoward enzalutamide-venetoclax interactions observed in our study and considering that new BCL-2 inhibitors in clinical trials such as lisaftoclax (APG-2575) (48) and sorotoclax (BGB-11417) (49,50) are also CYP3A4 substrates, future studies should explore the combination of a different ARPI such as abiraterone, which is known to be not a CYP3A4 inducer (but a CYP3A4 substrate), with venetoclax to optimize BCL-2 targeting and potentially improve treatment outcomes in mCRPC patients. Regardless, the combination of ARPIs and BCL-2 inhibition may likely delay the PCa resistance to ARPIs. By simultaneously targeting heterogeneous (AR^+^ and AR^−/lo^) and highly plastic PCa cell populations and metastatic foci, this approach could potentially prevent the emergence of resistant clones and prolong the durability of response to ARPIs.

## Authors’ contributions

S.P, D.G, R.J, C.J and G.C led the clinical trial; A.J, J.K and D.G.T were involved in correlative and mechanistic studies; J.L.H, R.A.P, R.B and J.H.B led the PK studies; K.W was responsible for clinical trial related statistics work; D.Q. provided venetoclax and financial support for the trial; S.P, A.J, J.S.K, J.H.B, X.L, D.G.T and G.C were involved in manuscript draft writing. D.G.T and G.C finalized manuscript writing.

## Acknowledgements

The clinical trial was supported by a grant from Abbvie and a Prostate Cancer Foundation (PCF) Challenge Award (2022CHAL3788) to G.C. The PK portion of this study was supported in part by NCI grant R50CA211241 and this project used the UPMC Hillman Cancer Center (HCC) Cancer Pharmacokinetics and Pharmacodynamics Facility (CPPF) and was supported in part by award P30CA047904 (J.H.B). Correlative and molecular studies were supported, in part, by grants from the U.S National Institutes of Health (NIH) National Cancer Institute (NCI) R01CA237027, 2R01CA240290-06 and R21CA237939, grants from the U.S Department of Defense (DOD) PC220137 and PC220273, and Roswell Park Comprehensive Cancer Center and the NCI Center grant P30CA016056 (D.G.T).

## Note

Supplementary data for this article are available at Clinical Cancer Research Online (http://clincancerres.aacrjournals.org/).

## SUpp

**Supplementary Table S1.**
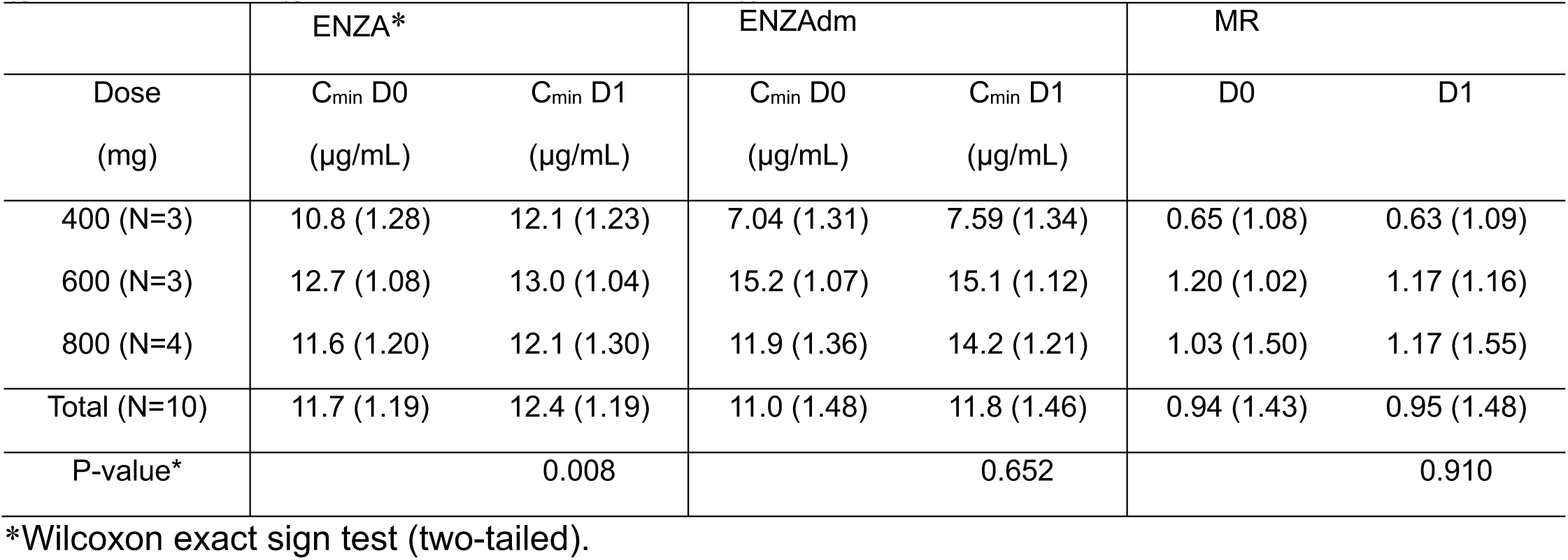
Enzalutamide and N-desmethyl enzalutamide plasma PK parameters (geometric mean (geometric standard deviation)).

**Supplementary Fig. S1.**
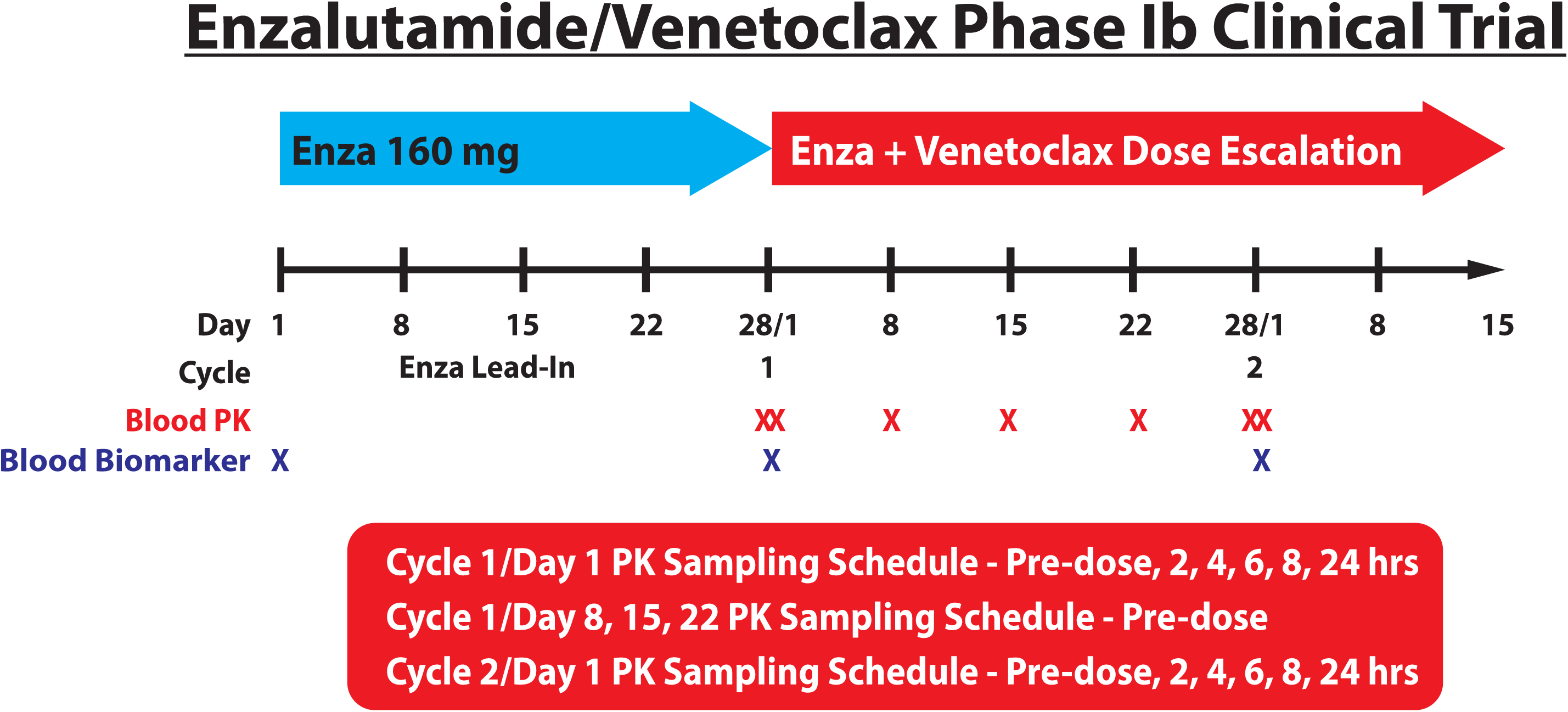
The phase Ib enzalutamide/venetoclax combination trial schema. See “Study design and objectives” in Methods for details.

**Supplementary Fig. S2.**
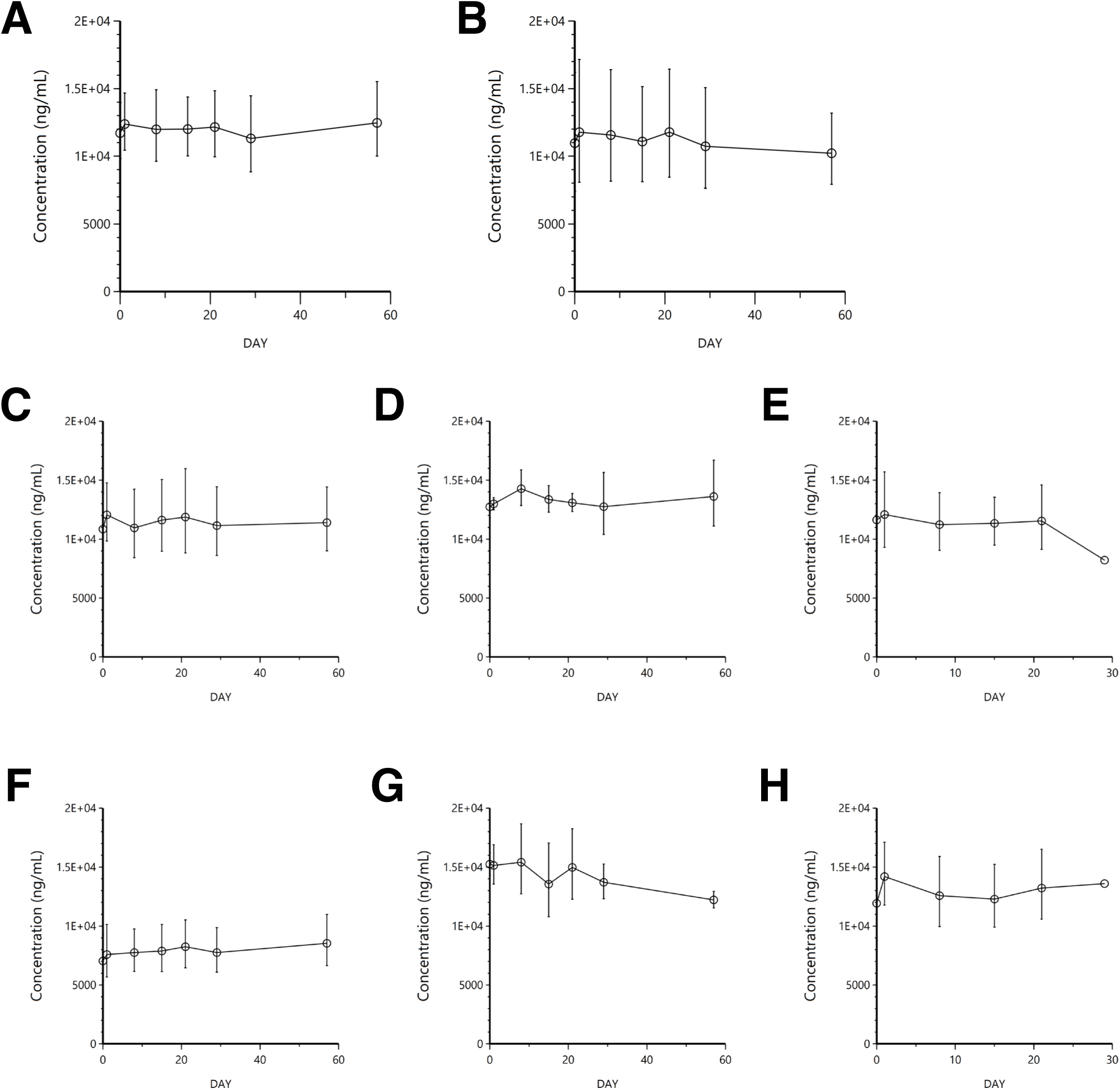
Enzalutamide PK studies. (**A-B**) Trough Enzalutamide (A) and N-desmethyl Enzalutamide (B) concentrations (geometric mean±geometric standard deviation) over the period of study with venetoclax

**Supplementary Fig. S3.**
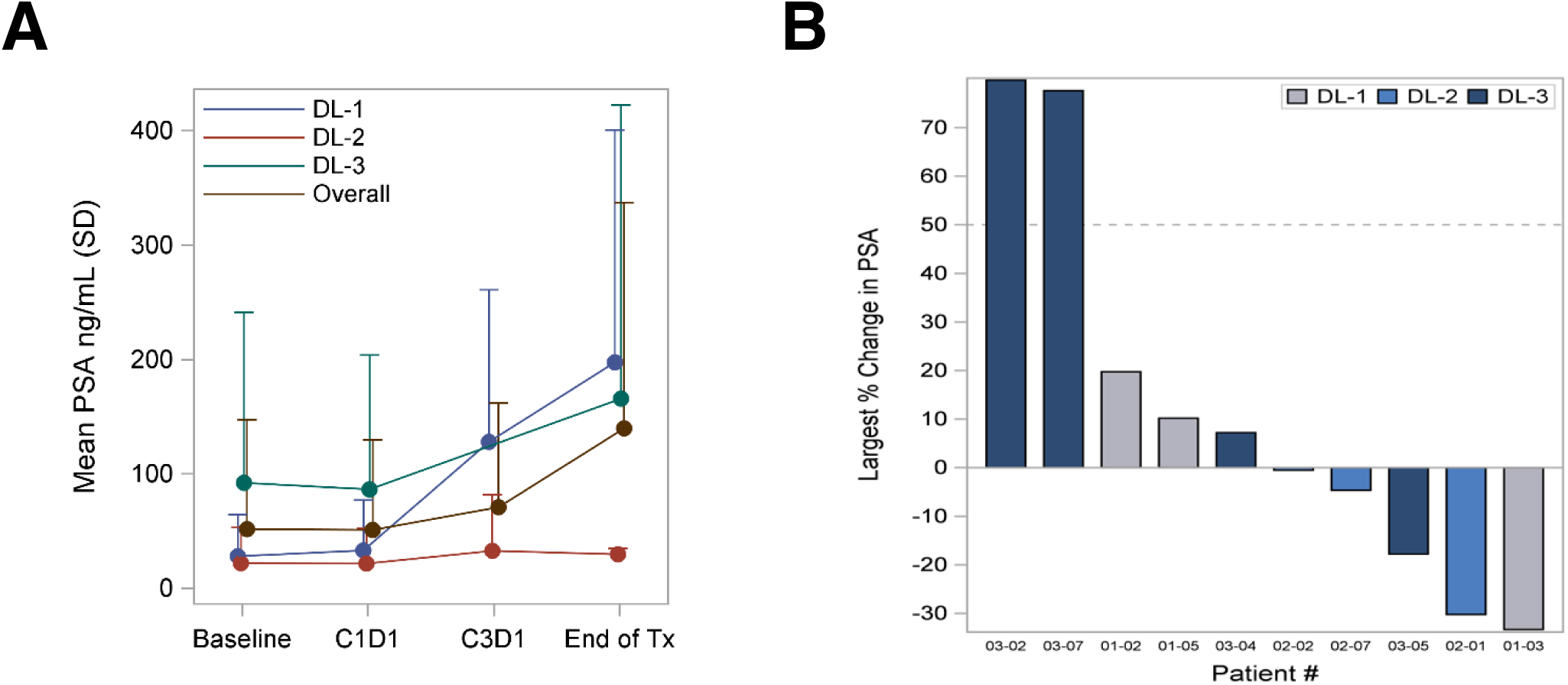
Measures of clinical efficacy of the enzalutamide/venetoclax combination treatment in mCRPC patients (**A-B**) Shown are PSA response summary (A) and waterfall plot (B).

